# Predicting Oncogene Mutations of Lung Cancer Using Deep Learning and Histopathologic Features on Whole-Slide Images

**DOI:** 10.1101/2022.05.03.22274614

**Authors:** Naofumi Tomita, Laura J. Tafe, Arief A. Suriawinata, Gregory J. Tsongalis, Mustafa Nasir-Moin, Konstantin Dragnev, Saeed Hassanpour

## Abstract

Lung cancer is a leading cause of death in both men and women globally. The recent development of tumor molecular profiling has opened opportunities for targeted therapies for lung adenocarcinoma (LUAD) patients. However, the lack of access to molecular profiling or cost and turnaround time associated with it could hinder oncologists’ willingness to order frequent molecular tests, limiting potential benefits from precision medicine. In this study, we developed a weakly supervised deep learning model for predicting somatic mutations of LUAD patients based on formalin-fixed paraffin-embedded (FFPE) whole-slide images (WSIs) using LUAD subtypes-related histological features and recent advances in computer vision. Our study was performed on a total of 747 hematoxylin and eosin (H&E) stained FFPE LUAD WSIs and the genetic mutation data of 232 patients who were treated at Dartmouth-Hitchcock Medical Center (DHMC). We developed our convolutional neural network-based models on 172 training cases and tested on 60 independent cases to analyze whole slides and predict five major genetic mutations, i.e., *BRAF, EGFR, KRAS, STK11*, and *TP53*. We additionally used 111 cases from the LUAD dataset of the CPTAC-3 study for external validation. Our model achieved an AUROC of 0.799 (95% CI: 0.686-0.904) and 0.686 (95% CI: 0.620-0.752) for predicting *EGFR* genetic mutations on the DHMC and CPTAC-3 test sets, respectively. Predicting *TP53* genetic mutations also showed promising outcomes. Our results demonstrated that H&E stained FFPE LUAD whole slides could be utilized to predict oncogene mutations, such as *EGFR*, indicating that somatic mutations could present subtle morphological characteristics in histology slides, where deep learning-based feature extractors can learn such latent information.

## Introduction

Lung cancer is a leading cause of death in both men and women in the world. In 2020, 1.8 million individuals have died from lung cancer, and 2.2 million cases are newly diagnosed (1). Non-small cell lung carcinoma (NSCLC) accounts for more than 80% of lung cancer cases, and lung adenocarcinoma (LUAD) is one of the most prevalent histologic subtypes of NSCLC. The recent development of molecular profiling has opened new targeted therapy opportunities for LUAD patients, which can improve clinical outcomes and the quality of life of patients. Several actionable mutations have been identified for LUAD targeted treatment: *KRAS, EGFR, ALK, MET, BRAF, RET, ROS1, NTRK*, and *ERBB2* (2, 3). The mutation frequencies and the clinical implications of each mutation in NSCLC patients vary. For example, *EGFR* somatic mutation is present in 12-15% of the Caucasian population with NSCLC and 47-64% of East Asian NSCLC patients. Its reported Overall Response Rate (ORR) to Osimertinib, a third-generation *EGFR* tyrosine kinase inhibitor (TKI), is about 80%. In contrast, the reported ORR of *KRAS p*.*G12C* mutation targeted drug, Sotorasib, is 32% (3).

Treatments targeting these mutations have improved the survival rate of NSCLC male patients from 26% in 2001 to 35% in 2014 (4). Next-generation sequencing (NGS) testing, which is performed on tumor tissue samples to identify somatic mutations, is used in the current standard of care for advanced NSCLC patients (5). In a recent survey in the United States, over 75% of oncologists use NGS tests to guide their treatment decisions for patients (6). This survey, however, also revealed the low frequency of ordering NGS testing. While the cause of the low frequency of NGS testing is not well studied, lack of access, long turnaround time (typically 10-14 days), and the cost of testing could hinder the oncologists’ willingness to order NGS testing. There are potentially more patients who could benefit from performing tumor molecular profiling to decide treatment eligibilities in precision oncology.

There has been a new interdisciplinary development at the intersection of clinical oncology and machine learning research to predict the actionable mutations in cancer tissues based on formalin-fixed paraffin-embedded (FFPE) whole-slide images without the need for molecular profiling tests for colorectal cancer (7), gastric cancer (8), breast cancer (9, 10) and lung cancer (11-13). Coudray et al. applied a convolutional neural network (CNN) to predict ten major mutations and demonstrated that predicting the mutation status of *STK11, EGFR, FAT1, SETBP1, KRAS*, and *TP53* is a feasible task (11). In this study, we set up a novel weakly supervised framework to train a deep learning model for patient-level somatic mutation prediction with validation on both internal (DHMC) and external (CPTAC-3) datasets, adding new methodological improvements and experimental evidence to the existing body of work to advance this field. We hypothesize that LUAD subtype-related histopathology features extracted using a CNN could be utilized to further predict the oncogene mutations. To this end, we develop a new weakly supervised deep learning model for predicting somatic mutations based on FFPE whole-slide images (WSIs) of LUAD patients. Ultimately, the successful development of such algorithms to identify oncogene mutations based on whole slide images would be a great benefit for both patients and healthcare systems by providing a triaging method that could be utilized before performing time-intensive and expensive molecular testing to screen patients for clinically actionable mutations and identify and prioritize those cases that likely benefit from targeted treatments in a more timely manner.

## Methods

### Datasets

A total of 747 hematoxylin and eosin (H&E) stained FFPE lung adenocarcinoma (LUAD) whole-slide images and their corresponding genomic profile were collected from 232 patients who were treated at the Dartmouth Hitchcock Medical Center between 2018 and 2019. These H&E stained slides were digitized by an Aperio AT2 scanner (Leica Biosystems, Wetzlar, Germany) at 40x magnification (0.25 µm/pixel). The genetic mutation data was generated by next generation sequencing (NGS) as part of routine patient treatment at DHMC. The NGS panel used for these samples covered hotspot mutation regions in 50 cancer related genes. These 50-gene hotspot regions cover the most relevant genetic information for precision-medicine lung-cancer management. These regions and their relevance to lung cancer were established through a rigorous independent research and selection process, which included a systematic review (by multiple domain expert genomic, pathology, and oncology researchers at DHMC and Norris Cotton Cancer Center) of the most prominent NSCLC knowledge bases, such as the National Comprehensive Cancer Network (NCCN) Clinical Practice Guidelines in Oncology, My Cancer Genome: Genetically Informed Cancer Medicine, COSMIC: Catalogue of Somatic Mutations in Cancer, ClinVar National Center for Biotechnology Information, dbSNP National Center for Biotechnology Information, and literature searches using PubMed (14-21). Curation of these hotspot regions and determination of their clinical importance are previously published and established (14-20).

We binarized the NGS data for these 50-gene hotspots to indicate whether a mutation for each gene was present. We considered five genes, *BRAF, EGFR, KRAS, STK11*, and *TP53*, in this study because those were the genes that were mutated in at least five percent of all patients in our dataset. Three oncogenes, *BRAF, EGFR* and *KRAS*, are typically mutually exclusive. However, we identified two cases in our dataset with mutations in both *BRAF* and *KRAS*. This overlap has also been reported in previous studies on these mutations in NSCLC patients (22). Of note, two tumor suppressor genes, *TP53* and *STK11*, can overlap with the oncogenes, particularly *TP53*. In our dataset, we found 24 co-occurrences of mutations in *KRAS* and *TP53*, 7 co-occurrences of mutations in *BRAF* and *TP53*, 6 co-occurrences of mutations in *KRAS* and *STK11*, 4 co-occurrences of mutations in *KRAS, STK11*, and *TP53*, 2 co-occurrences of mutations in *BRAF, KRAS*, and *TP53*, and 1 co-occurrence of mutations in *EGFR* and *STK11*. We randomly partitioned the slides stratified by patient into train, validation, and test set, containing 471, 97, and 179 cases, respectively. Due to the heterogeneous distribution of genetic mutations in our dataset, we ensured each partition included at least one patient with mutation for these five genes. Table 1 summarizes the distribution of patients and their genetic mutation status in our dataset.

**Table 1.**
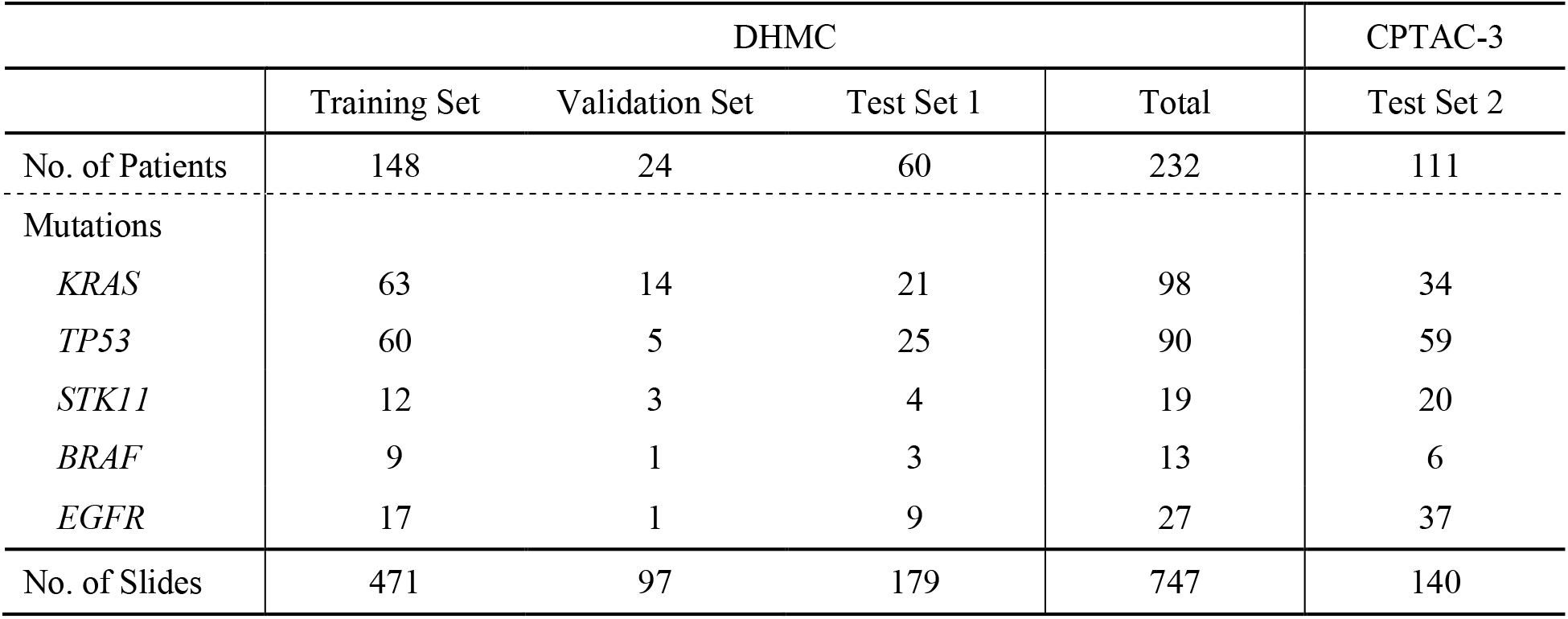
The distribution of patients and their mutation status in our datasets.

Additionally, we collected 140 H&E stained FFPE slides and corresponding genetic mutation data of 111 lung cancer patients from the Clinical Proteomic Tumor Analysis Consortium 3 (CPTAC-3), as external validation (23). This study and the use of human participant data in this project were approved by the Dartmouth-Hitchcock Health Institutional Review Board (IRB) with a waiver of informed consent. The conducted research reported in this study is in accordance with this approved Dartmouth-Hitchcock Health IRB protocol and the World Medical Association Declaration of Helsinki on Ethical Principles for Medical Research involving Human Subjects.

### Data Preprocessing

Since digitized slides consist of multi-million pixels, which current common computational hardware cannot easily process at once, we preprocessed each whole-slide image in our dataset and extracted smaller fixed-size patches for our analysis. For this preprocessing, we first down-sampled each whole-slide image by a factor of eight (i.e., converted the images to 5x magnification or 2.0 µm/pixel), removed background and artifacts, and generated patches of 224×224 pixels using a sliding window approach (24) with an overlapping factor of 1/3 from these down-sampled whole-slide images.

We applied patch filtering with a CNN model pre-trained on a LUAD dataset for histologic subtype classification task (24) to remove an overwhelming number of patches with normal tissue and focused on LUAD-related regions of the whole slides. For this filtering, we applied the aforementioned pre-trained model (24) to predict the histological subtype of the patch (i.e., acinar, lepidic, micropapillary, papillary, solid, or normal), and removed patches predicted as normal. The patch filtering method removes normal patches that have less to no information about the tumor and effectively accelerates the model training by reducing the number of training samples. In addition, since we used a weakly supervised framework for CNN model training, removing normal patches was an essential preprocessing step for noise reduction.

### Deep Neural Network based Models

We took a bottom-up approach to analyze whole-slide images, where a set of fixed-size (i.e., 224×224 pixels) tissue patches from a digitized slide are fed to a CNN-based image feature extractor. Extracted features are aggregated and analyzed to predict the genetic mutation for a patient. To achieve an accurate whole-slide-based prediction of genetic mutation, we considered two types of image features: 1) LUAD subtype-specific features and 2) generic image features. The first features are extracted using a CNN model (i.e., ResNet18) that was trained for classifying LUAD subtypes using the DHMC LUAD subtypes dataset (24). The second features are based on an ImageNet-pretrained CNN model, which is implemented in one of the state-of-the-art CNN architectures (i.e., EfficientNetB0 architecture)(25). In the deep learning paradigm, it is commonly preferred to use a feature extractor that is pre-trained on a relevant task in the same modality; however, there is also an increasing number of reports in the biomedical domain that shows general feature extractors could be beneficial with minimal fine-tuning (9). This is because the ImageNet dataset is usually far larger than medical image datasets in terms of sample size and target classes, which could unlock its generic power to extract subtle features even in a different modality. We explored different model architectures in this work to examine the efficacy of reusing an off-the-shelf pathology-specific feature extractor and also off-the-shelf generic feature extractor. In this study, CNN_LUAD-Feat_ denotes a CNN model pretrained on the LUAD subtype classification task, and CNN_Image-Feat_ denotes a CNN pretrained on the ImageNet classification task. Both models were fine-tuned for our patch-level genetic mutation prediction task at two levels: 1) only the last fully-connected layer or 2) all the layers except for batch normalization layers. For patient-level genetic mutation prediction, all the patch-level predictions are pooled and aggregated to compute the average confidence score for each somatic mutation. We applied a grid-search optimization on the average confidence score in the validation set to establish our confidence score threshold for patient-level inference.

In addition to these two CNN models, we developed a LUAD subtype distribution-based method to investigate the translational power of LUAD subtype predictions to genetic mutation prediction. Instead of using the LUAD imaging features, we reused the LUAD subtype classification results of tissue patches and the proportion of each LUAD subtype area on whole slides. To this end, we trained a logistic regression model that takes a slide-level LUAD subtype distribution for a patient as input and predicts somatic mutation status for each gene. In this work, Logit_LUAD-hist_ denotes the logistic regression model that was developed on top of the pretrained CNN for LUAD subtype predictions. Figure 1 shows the overview of our approaches in this study.

**Figure 1.**
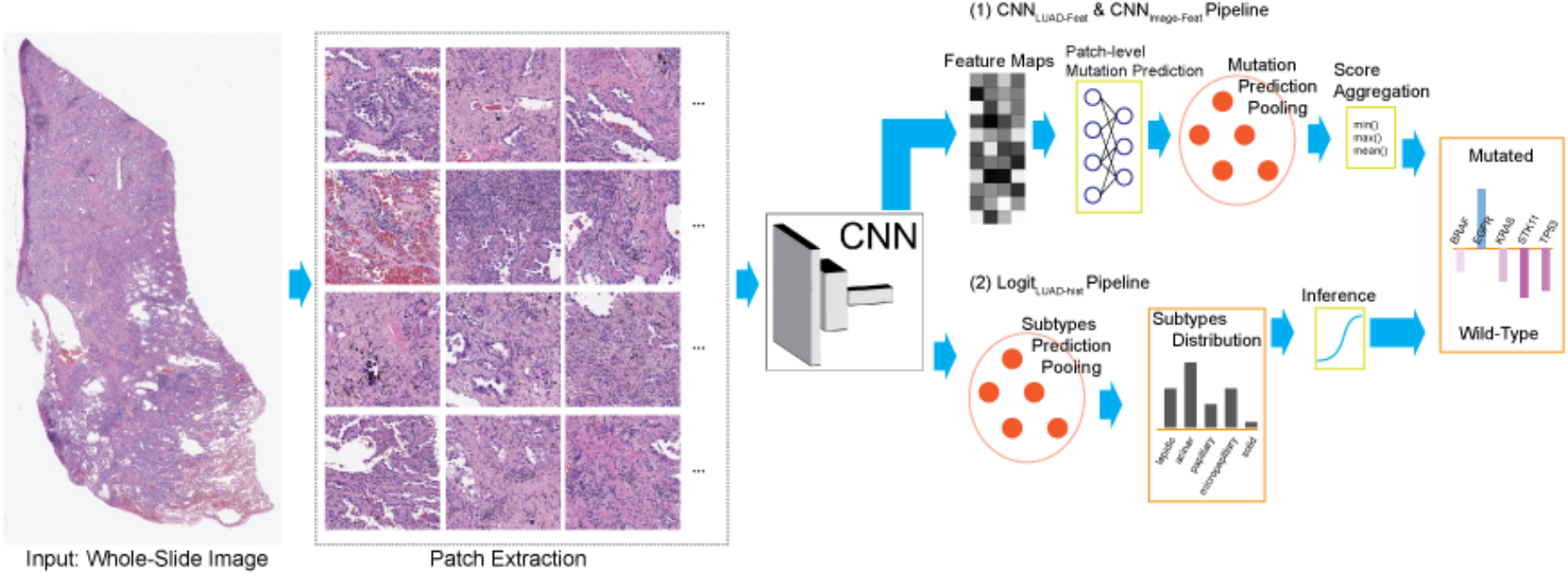
Overview of our pipelines. Tissue patches are extracted from whole-slide images using a sliding-window method with one-third overlap after removing background. (1) Extracted features through a CNN are used to predict patch-level mutation level. The predictions are pooled and aggregated to compute a confidence score for each somatic mutation. (2) LUAD subtype classification results of patches are pooled to compute the proportion of each LUAD subtype area on whole slides. A logistic regression is applied to the LUAD subtype distribution to predict somatic mutation status for each gene.

### Evaluation Metrics and Statistical Analysis

We evaluated our method on two different test sets: 1) an internal test set of 60 patients from DHMC and 2) an external test of 111 patients from the CPTAC-3 dataset to show the generalizability of our approach. Each genetic mutation was independently predicted (i.e., five binary classifications for each slide), and we used the area under the receiver operating characteristics (AUC) for each genetic mutation prediction to evaluate the performance of our models. In addition, we computed 95% confidence intervals (95% CIs) using the bootstrapping method with 1,000 iterations for each metric.

## Results

Table 2 summarizes the evaluation results of our CNN models on the internal test set from DHMC. Each model name is followed by either “-FT/FC” or “-FT/AL”, where “-FT/FC” indicates the model is fine-tuned at the last fully-connected layer, and “-FT/AL” indicates the model is fine-tuned at all of the layers (except batch-normalization layers). The CNN_LUAD-Feat- FT/FC_ model on the first row of this table achieved an AUC of 0.804 (95% CI: 0.614-0.972) for *BRAF* mutation and an AUC of 0.711 (95% CI: 0.616-0.803) for *TP53* mutation. The CNN_Image- Feat-FT/AL_ on the fourth row achieved an AUC of 0.799 (95% CI: 0.686-0.904) for *EGFR* mutation and an AUC of 0.713 (95% CI: 0.611-0.811) for *TP53* mutation.

**Table 2.** AUCs and associated 95% CI achieved by our models on the internal DHMC test set for each somatic mutation. A model name followed by “-FT/FC” indicates the model is fine-tuned at the last fully-connected layer. A model name followed by “-FT/AL” indicates the model is fine-tuned at all of the layers (except batch-normalization layers). The AUC of 0.65 or higher is highlighted in bold.

| Models | *BRAF* | *EGFR* | *KRAS* | *STK11* | *TP53* |
| --- | --- | --- | --- | --- | --- |
| $\text{CNN}_{\text{LUAD-Feat-FT/FC}}$ | **0.804**<br>(**0.614-0.972**) | 0.627<br>(0.483-0.764) | 0.604<br>(0.490-0.719) | 0.612<br>(0.407-0.800) | **0.711**<br>(**0.616-0.803**) |
| $\text{CNN}_{\text{LUAD-Feat-FT/AL}}$ | 0.483<br>(0.197-0.767) | 0.570<br>(0.446-0.687) | 0.530<br>(0.419-0.639) | 0.606<br>(0.392-0.802) | 0.635<br>(0.528-0.741) |
| $\text{CNN}_{\text{Image-Feat-FT/FC}}$ | 0.518<br>(0.244-0.780) | 0.479<br>(0.348-0.605) | 0.474<br>(0.364-0.582) | 0.521<br>(0.262-0.769) | 0.565<br>(0.456-0.672) |
| $\text{CNN}_{\text{Image-Feat-FT/AL}}$ | 0.543<br>(0.297-0.774) | **0.799**<br>(**0.686-0.904**) | 0.596<br>(0.486-0.706) | 0.609<br>(0.374-0.832) | **0.713**<br>(**0.611-0.811**) |

Table 3 summarizes the evaluation of our CNN models on the external CPTAC-3 test set. CNN_Image-Feat-FT/AL_ achieved an AUC of 0.686 (95% CI: 0.620-0.752) for *EGFR* mutation and an AUC of 0.677 (95% CI: 0.602-0.752) for *TP53*, showing a consistent performance across different datasets. On the contrary, the CNN_LUAD-Feat-FT/FC_, which had high predicting performance for *BRAF* and *TP53* mutations on the internal DHMC test set, did not achieve a consistent performance on the external CPTAC-3 test set.

**Table 3.**
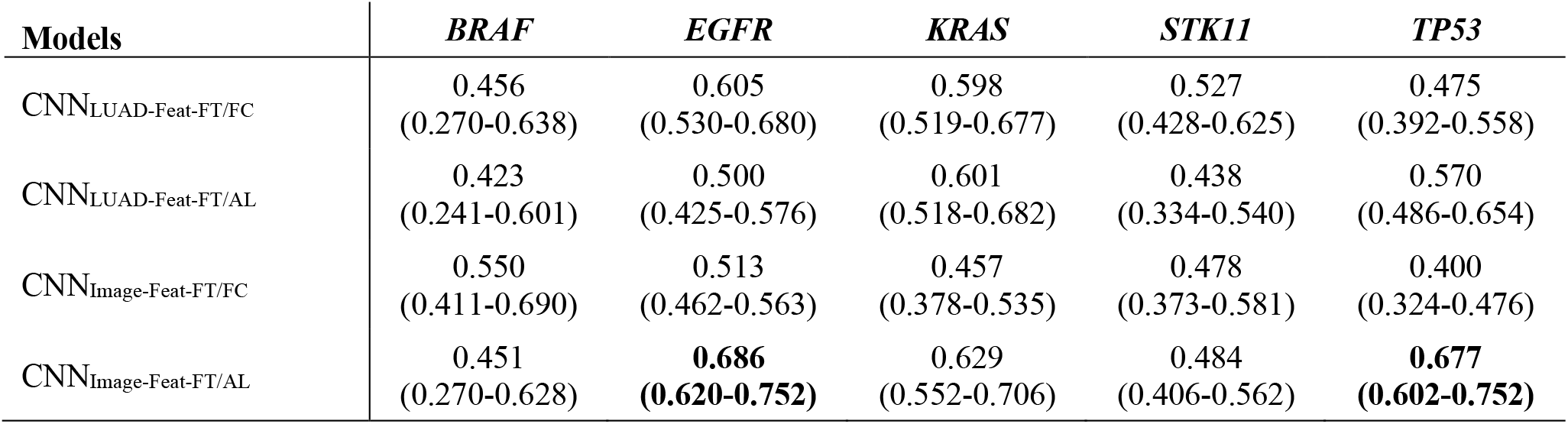
AUC with 95% CI achieved by our models on the external CPTAC-3 test set for each somatic mutation. A model name followed by “-FT/FC” indicates the model is fine-tuned at the last fully-connected layer. A model name followed by “-FT/AL” indicates the model is fine-tuned at all of the layers (except batch-normalization layers). The AUC of 0.65 or higher is highlighted in bold.

Figure 2 illustrates the receiver operating characteristics (ROC) curves of CNN_Image-Feat- FT/AL_ model for each oncogene mutation across the DHMC and CPTAC-3 test sets.

**Figure 2.**
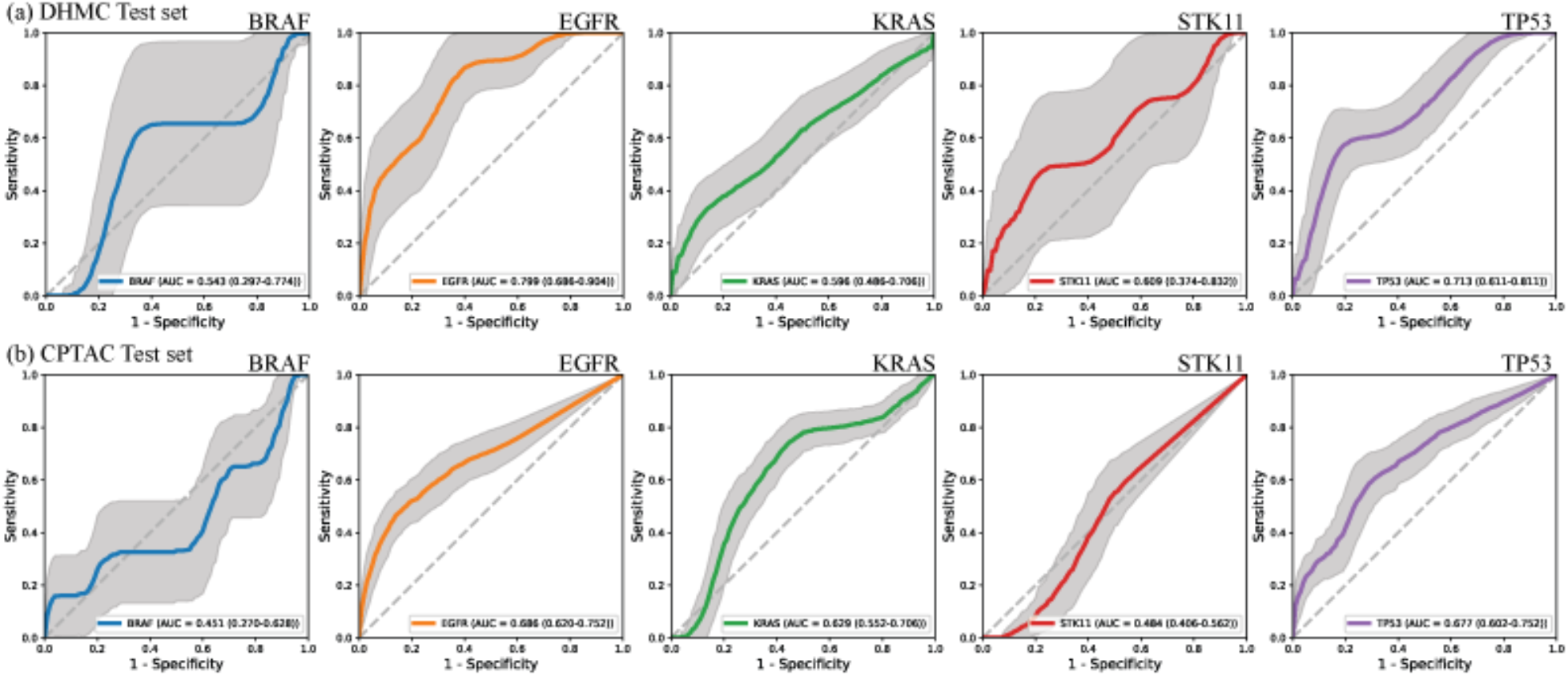
The receiver operating characteristic curves (ROC) of CNN_Image-Feat-FT/AL_ for each somatic mutation predicted for (a) internal DHMC test set and (b) external CPTAC-3 test set. Each column from left to right corresponds to *BRAF, EGFR, KRAS, STK11*, and *TP53*, respectively. The gray bands represented 95% confidence intervals of each ROC.

Table 4 shows the performance of our logistic regression model built on top of the pretrained CNN’s LUAD subtype distribution. Logit_LUAD-hist-mean_, which employs the mean aggregation for slide-based LUAD subtype distributions of a patient, achieved an AUC of 0.681 (95% CI: 0.567-0.770) for *EGFR* mutation and an AUC of 0.692 (95% CI: 0.580-0.778) for *TP53* mutation on the internal DHMC test set. Logit_LUAD-hist-min_, which similarly used the minimum aggregation, achieved an AUC of 0.725 (95% CI: 0.623-0.803) for *BRAF* mutation and an AUC of 0.69 (95% CI: 0.578-0.777) for *TP53* mutation on the internal DHMC test set. The models, however, did not perform better than an AUC of 0.6 on the external CPTAC-3 test set.

**Table 4.** AUC with 95% CI achieved by our logistic regression models based on LUAD subtype distribution on internal DHMC and external CPTAC-3 test sets for each somatic mutation. A model name followed by “-max”, “-mean”, or “-min” indicates the model uses the max, mean, and min function, respectively, as aggregation of slide-level predictions for a patient. The AUC of 0.65 or higher is highlighted in bold.

| Datasets | Models | *BRAF* | *EGFR* | *KRAS* | *STK11* | *TP53* |
| --- | --- | --- | --- | --- | --- | --- |
| DHMC | Logit<sub>LUAD-hist-max</sub> | 0.456<br>(0.295-0.592) | **0.66**<br>**(0.540-0.754)** | 0.506<br>(0.353-0.633) | 0.5<br>(0.346-0.628) | **0.661**<br>**(0.541-0.754)** |
|  | Logit<sub>LUAD-hist-mean</sub> | 0.578<br>(0.439-0.690) | **0.681**<br>**(0.567-0.770)** | 0.515<br>(0.364-0.640) | 0.5<br>(0.346-0.628) | **0.692**<br>**(0.580-0.778)** |
|  | Logit<sub>LUAD-hist-min</sub> | **0.725**<br>**(0.623-0.803)** | 0.631<br>(0.504-0.731) | 0.5<br>(0.346-0.628) | 0.5<br>(0.346-0.628) | 0.69<br>(0.578-0.777) |
| CPTAC-3 | Logit<sub>LUAD-hist-max</sub> | 0.158<br>(0.0-0.335) | 0.468<br>(0.309-0.602) | 0.553<br>(0.409-0.670) | 0.5<br>(0.346-0.628) | 0.576<br>(0.436-0.688) |
|  | Logit<sub>LUAD-hist-mean</sub> | 0.163<br>(0.0-0.339) | 0.475<br>(0.317-0.608) | 0.562<br>(0.420-0.677) | 0.5<br>(0.346-0.628) | 0.578<br>(0.439-0.690) |
|  | Logit<sub>LUAD-hist-min</sub> | 0.186<br>(0.0-0.360) | 0.488<br>(0.332-0.618) | 0.57<br>(0.429-0.684) | 0.5<br>(0.346-0.628) | 0.583<br>(0.445-0.694) |

## Discussion

In this study, we developed several deep neural network-based models that predict oncogene mutations based on H&E stained FFPE whole-slide images of LUAD patients to investigate the utilization of existing pretrained models that were developed for a different task. Our experiments showed promising performance in predicting *EGFR* and *TP53* mutations, achieving an AUC of 0.799 and 0.713 on the internal DHMC test set, using an ImageNet pretrained CNN model with fine-tuning all the layers. A model that fine-tuned only the last fully-connected layer and directly reused the generic image-based features did not achieve the best performance, indicating that ImageNet-based generic image features might not be directly applicable to our task. A model with ImageNet parameters, however, could be a good starting point to fine-tune for further domain-specific tasks, confirming a common ground in deep learning research (26, 27). We also observed that a model using LUAD features had limited generalizability in predicting *BRAF* and *TP53* mutations when tested on the external test set. A similar trend was observed in our experiment with logistic regression models using LUAD subtype distribution where prediction performance for *BRAF, EGFR*, and *TP53* mutations was declined in the external test set. While the LUAD subtype dataset for pretraining a CNN model and the LUAD dataset in this study were collected independently, we hypothesize that there might exist some internal consistency, such as tissue preparation or scanner type, that could lead to models’ overfitting.

Of note, *EGFR* mutations, which one of our proposed models identifies at an AUC of 0.799 based on whole-slide images in the DHMC test set, is an important factor in the targeted treatment of NSCLC patients. Currently, Osimertinib, an *EGFR* inhibitor, is approved by the US Food and Drug Administration (FDA) for the treatment of NSCLC with stage IB and above and have led to improvements in clinical outcome and quality of life of NSCLC patients with *EGFR*-mutation (28, 29).

Any delay in genetic testing of NSCLC patients with potential clinically-actionable mutations, such as *EGFR*, can have major impacts on patients clinical outcomes. We expect further development and validation of the presented methods in this work could lead to new approaches to identify NSCLC patients with clinically-actionable mutations based on tumor pathology slides, and to provide an accurate, fast, and inexpensive pre-selection method that could be utilized before performing time-intensive and expensive genetic tests to screen patients for clinically-actionable mutations. As a result, these prediction methods could prioritize genetic screening of NSCLC patients who are the most likely to have clinically-actionable mutations, thus reducing screening turnaround time and increasing the accuracy of treatment administration. In addition, such pre-selection methods could improve the finding and tracking of NSCLC patients with clinically-actionable mutations for translational research, as well as facilitate the recruitment of NSCLC patients for clinical trials.

Our study further supports oncogene mutation prediction using deep learning with both internal and external test sets, suggesting that gene mutations could present subtle morphological characteristics in whole slides, where deep learning-based feature learners can extract such latent information. Of note, utilizing histopathology features of LUAD subtypes had limited utility in predicting oncogene mutations. Still, our experiments showed promising results for predicting *BRAF, EGFR*, and *TP53* mutations based whole-slide image features. As a future direction, we plan to investigate *KRAS* and *STK11* mutations with alternative approaches. In addition, we plan to extend our histopathology-based analysis to further predict response levels and time to the development of resistance for targeted therapies.

## Data Availability

CPTAC-3 data can be downloaded from the website: https://portal.gdc.cancer.gov/projects/CPTAC-3. The DHMC dataset used in this study is not publicly available due to patient privacy constraints. An anonymized version of this dataset can be generated and shared upon reasonable request from the corresponding author.

## Contributors

Concept and design: L.T. and S.H.; Acquisition, analysis, or interpretation of data: All authors; Drafting of the manuscript: M.N., N.T., and S.H.; Critical revision of the manuscript for important intellectual content: All authors.; Statistical analysis: M.N. and N.T.; Obtained funding: S.H.; Administrative, technical, or material support: S.H.; Supervision: S.H.

## Acknowledgements

This research was supported in part by grants from the US National Cancer Institute (R01CA249758) and the US National Library of Medicine (R01LM012837). The authors wish to acknowledge the support of the Pathology Shared Resource in the Laboratory for Clinical Genomics and Advanced Technology of the Dartmouth-Hitchcock Health System and the Norris Cotton Cancer Center at Dartmouth with NCI Cancer Center Support Grant 5P30 CA023108-37. The funders had no role in study design, data collection, data analysis, interpretation, decision to publish, or preparation of this manuscript.

